# Limitations of Public Biomechanical Movement Datasets for Deep Learning: Issues of Metadata, Standardization, and Variety in Motion Types

**DOI:** 10.1101/2025.05.29.25328474

**Authors:** Daniel Friemert, David Schnur, Simon Runkel, Jonas Borsch, Kiros Karamanidis, Babette Dellen, Lutz Thieme, Armin Fiedler, Uwe Jaekel, Ulrich Hartmann

**Affiliations:** University of Applied Sciences Koblenz, Joseph-Rovan-Allee 2, Remagen, Germany, 53424; Institute for Medical Technology and Information Processing, Universitätsstraße 1, Koblenz, Germany, 56070; Institute for Sports Management and Sports Medicine Technology, Joseph-Rovan-Allee 2, Remagen, Germany, 53424; London South Bank University, School of Applied and Health Sciences, 103 Borough Road, London, SE10AA, United Kingdom; Department of Sport Science, Faculty of Mathematics and Natural Sciences, University of Koblenz, Universitätsstraße 1, Koblenz, Germany, 56070

**Keywords:** Biomechanical Datasets, Human Activity Recognition, Artificial Intelligence, Machine Learning, Gait Analysis, Public Databases, Sensor Data, Motion Analysis

## Abstract

Biomechanical data play a crucial role in health research by providing relevant information about musculoskeletal mechanical loading and neuromuscular dysfunction during movement, which are essential for optimizing treatment strategies and improving patient outcomes. The relevance of such data for decision making processes in clinical settings depends on its quality and volume, as larger datasets enable more robust conclusions across diverse populations and conditions. In machine learning, extensive and representative datasets are vital for training algorithms to develop accurate predictive models. This systematic literature review explores the current landscape of biomechanical datasets and databases, examining existing resources, identifying gaps in data availability and quality, and discussing the potential for a new, structured database to address these shortcomings. We conducted a comprehensive search across PubMed, IEEE Xplore, Scopus, ScienceDirect, Springer, and arXiv using keywords related to biomechanics, gait, and human activity recognition (HAR). The review identified studies covering various motion types, data formats, and metadata availability. Our findings highlight significant challenges, including inconsistent metadata, lack of standardization during data acquisition and data processing, limited motion variety, and accessibility issues. Addressing these challenges through standardized data formats, comprehensive metadata, and enhanced accessibility could greatly advance decision making processes via machine learning approaches for clinical and therapeutic settings. This review emphasizes the need for improved biomechanical data resources to support the development of accurate machine learning models and innovative clinical interventions.

## 1 Introduction

Biomechanical data is becoming increasingly important in health research, providing accurate measurements of human movement that are essential for optimizing treatment strategies.^1^ The accurate representation of human movement enables health care professionals to deliver treatments more effectively, potentially improving patient outcomes. However, the value of such data depends not only on its quality, but also on the volume of available datasets. Larger datasets facilitate more robust and valid conclusions by providing a more comprehensive view of human biomechanics across diverse populations and conditions.^2^ When it comes to machine learning it is even more important that these datasets represent every facet of the examined population. Machine learning algorithms require extensive amounts of data for effective training, making feature richness of data critical to the development of accurate predictive models.^3^ In addition, the quality and quantity of biomechanical data is enhanced by the inclusion of metadata related to participants or study context. This metadata enables deeper insights and can improve the performance and outcomes of analytical algorithms by providing essential contextual information that supports more nuanced interpretations and applications.

This systematic literature review aims to provide a comprehensive overview of the current state of biomechanical datasets and databases in human movement research. It will inspect existing resources, identify potential gaps in data availability and quality, and discuss how a new, well-structured database could address these limitations. In doing so, this review will explore and provide helpful suggestions on how biomechanical data could be structured in the future. This will allow the potential of machine learning and big data analysis to be harnessed at full potential, eventually unlocking techniques for the field of biomechanics that have been inaccessible until now. The remainder of this paper is structured as follows: Section 2 describes the methods used to identify existing datasets. Section 3 presents the results of our search. Section 4 provides a detailed discussion of these results. Finally Section 5 concludes with a summary of our findings.hors on the journal homepage.

## 2 Methods

To capture a comprehensive set of publications relevant to biomechanical datasets and databases, we searched the following electronic databases: PubMed, IEEE Xplore, Scopus, ScienceDirect, Springer, and arXiv. These platforms were chosen for their extensive coverage of biomedical, engineering and scientific literature. The search strategy employed combinations of the following keywords: “Biomechanic” and “Dataset”, “Gait” and “Dataset”, “Biomechanic” and “Database”, “Gait” and “Database”, “HAR” and “Database”, “HAR” and “Dataset”.

### 2.1 Study Selection

The study selection process was structured into three distinct phases to ensure a thorough review. The first stage involved the screening of titles and abstracts. Two independent reviewers performed this task to determine the relevance of each article based on predefined inclusion and exclusion criteria. This initial filter helped to narrow down the number of potential studies for detailed review.^4^

After the initial screening, we performed a backward reference search, in which we examined the reference lists of all studies that passed the first stage to identify further relevant publications. This technique is particularly valuable for uncovering studies that may be cited in multiple sources but were missed in the initial database search.^5^

At the same time, a manual search was carried out to supplement the results from the electronic databases. This search involved using Google Scholar and other relevant online portals to search for gray literature and additional academic papers that may not be indexed in the main databases. This step was essential to ensure that no important studies were overlooked and that the dataset for the review was as comprehensive as possible.

## 3 Results

The initial search yielded 1148 records, with 488 records remaining after duplicates were removed (Figure 1). Following the manual research, 542 records were identified as requiring further analysis. Following the initial screening process, 80 full-text articles were subjected to a further assessment to determine their eligibility. The available scientific literature on this topic was limited, and only 45 articles met the extended inclusion criteria. The study characteristics include the following: motion type, data type, database or repository, and availability of the data. The overall quality of the data and the accompanying information is very poor. The specific challenges encountered with the articles deemed relevant for this study will be explained in the subsequent section.

**Fig 1.**
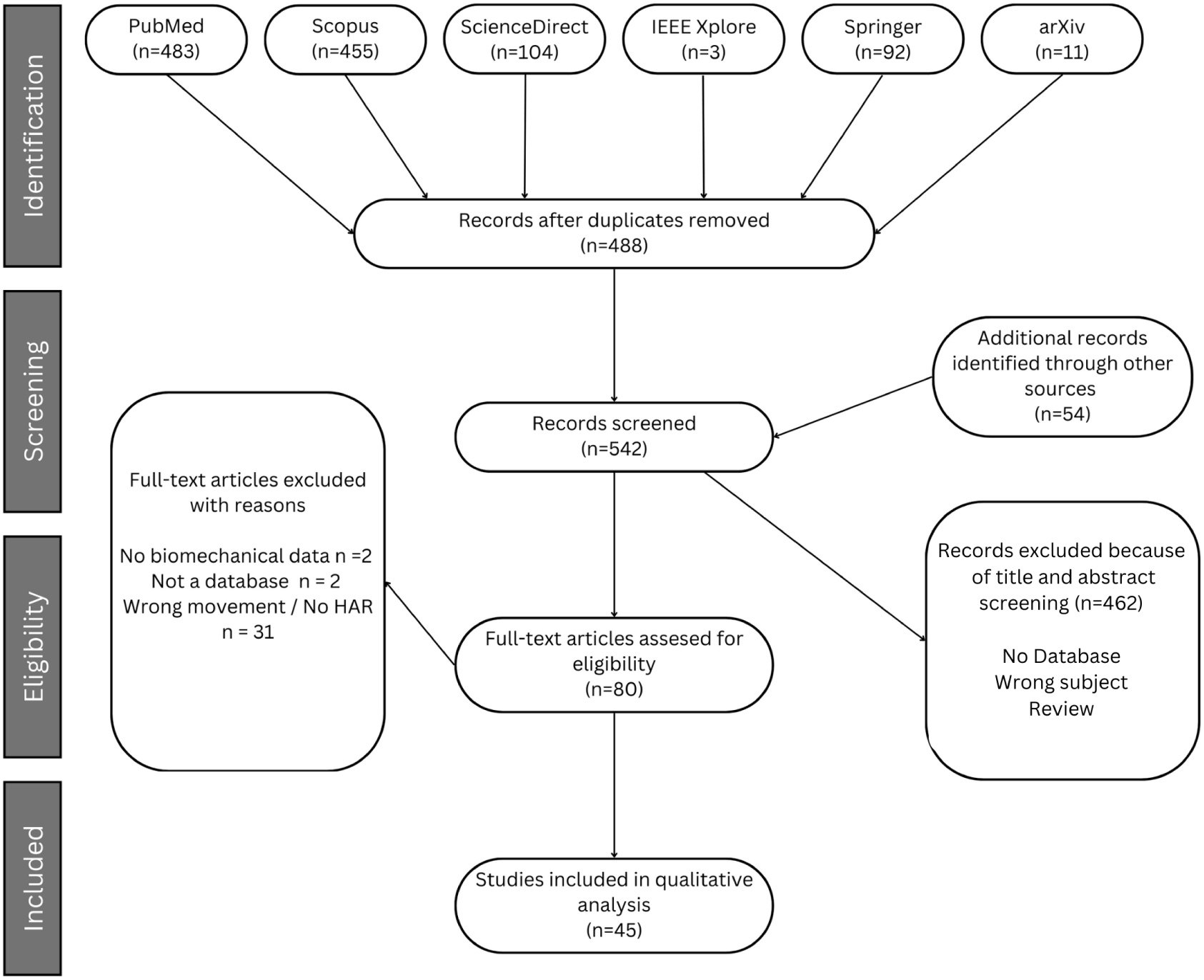
PRISMA flow diagram outlining the systematic review process for identifying, screening, and including studies on public biomechanics datasets for artificial intelligence. The diagram details the number of records retrieved from various databases, the screening and eligibility assessment steps, and the reasons for exclusion at each stage, ultimately resulting in the inclusion of 45 studies for qualitative analysis.

**Table 1.**
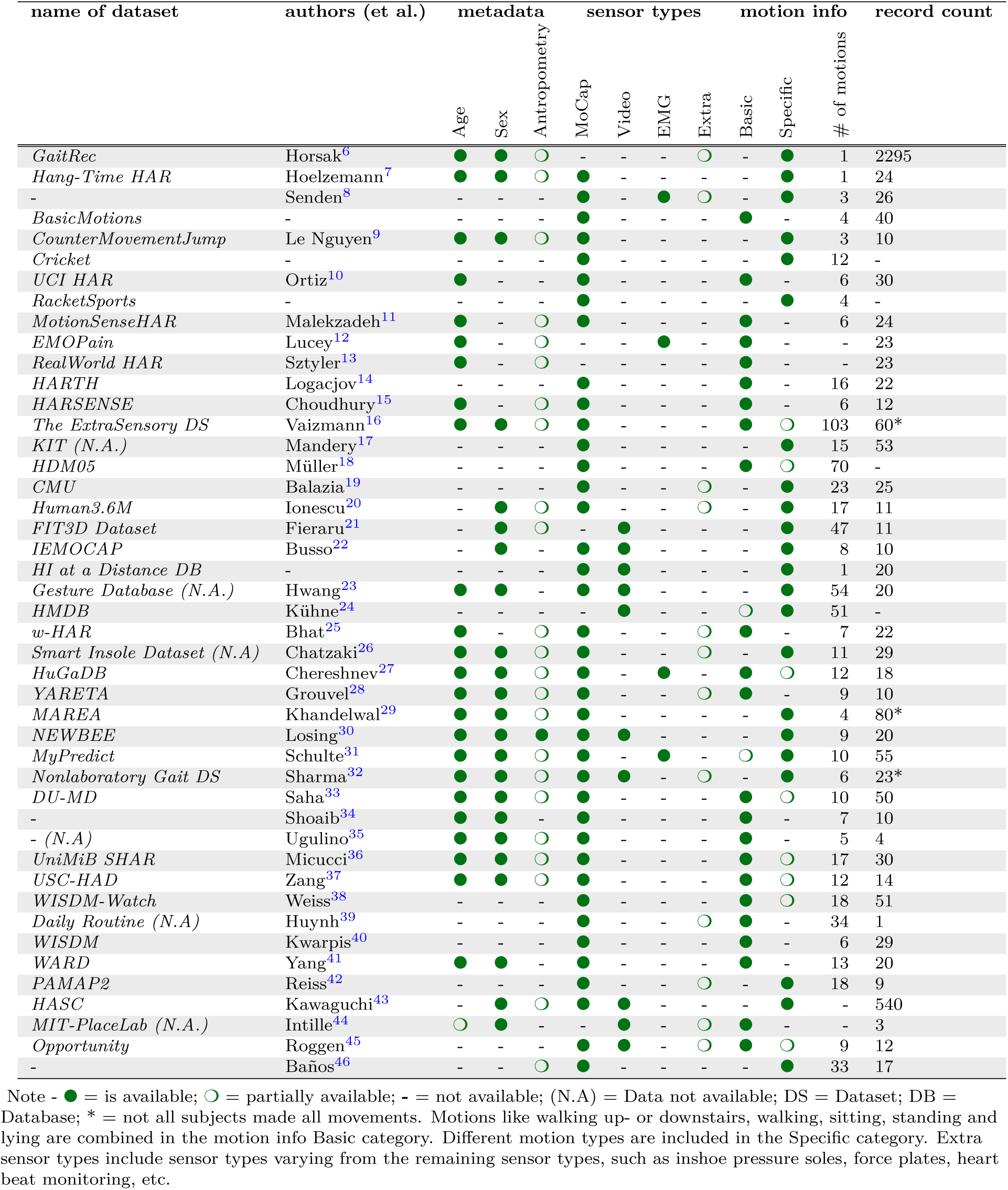
Results of the SLR for data records in motion analysis evaluated in the different categories of existing metadata, used sensor types, captured motions and motion variability and dataset volume.

This systematic literature review identified a multitude of databases and publicly accessible datasets pertinent to the field of human activity recognition. The key findings and characteristics of the reviewed datasets are presented in the following summary.

One of the most prominent datasets is GaitRec,^6^ described in the publication “GaitRec: A comprehensive ground reaction force dataset encompassing normal and pathological gait”. The dataset includes comprehensive metadata—such as gender, age, height, body weight, body mass, and shoe size—and provides is available in both CSV and Python script formats. Accessible via Figshare, an online digital repository, GaitRec offers valuable information for studying gait under both normal and pathological conditions. One notable advantage of this dataset is the comprehensive metadata recording, which facilitates in-depth analysis.

Another notable dataset is Hang-Time HAR,^7^ which was specifically designed for the purpose of basketball activity recognition. The publication “Hang-Time HAR: A Benchmark Dataset for Basketball Activity Recognition” provides a detailed description of this dataset, which includes a range of metadata—such as age, dominant hand, height, weight, and gender—and is available in CSV, TXT, and Python script. It is accessible via Zenodo, another open-access repository, with 25 records included. This dataset provides a specialized foundation for the development and testing of models for basketball activity recognition.

Another dataset by Senden and colleagues^8^ is described in the publication “Dataset of 3D Gait Analysis in Typically Developing Children and Young Adults”. Although this dataset lacks metadata, it offers various formats, including XLSX, JPG, PNG, and TXT, and is accessible via OSF, an open platform collaboration. With 26 records, this dataset is focused on 3D gait analysis in children and young adults, which can be relevant for specific studies but is limited by the absence of metadata.

The BasicMotions^47^ dataset is a time series classification dataset developed as part of a student project. It includes data from four students performing four different activities: walking, resting, running, and playing badminton. The data were captured using a smartwatch that recorded both 3D accelerometer and 3D gyroscope measurements. Each activity was recorded five times for 10 seconds per trial, with data sampled every 0.1 seconds. The dataset contains 40 training and 40 testing samples, and each time series has six dimensions that correspond to the three axes of both the accelerometer and gyroscope. This dataset is widely used to evaluate machine learning models for human activity recognition (HAR), especially focusing on time series data.

The CounterMovementJump (CMJ) dataset, contributed by Thach Le Nguyen,^9^ consists of multivariate time series data collected from participants performing a Counter Movement Jump (CMJ) test. The data captures three distinct classes of jumps: those performed with acceptable form, jumps with leg bending during flight, and jumps where participants stumbled upon landing. The dataset includes accelerometer data from the X, Y, and Z axes recorded using a Shimmer 3 Inertial Measurement Unit (IMU) attached to the dominant foot of each participant. Data was sampled at a high rate of 1024Hz, providing detailed motion data for each jump.

The UCI HAR Dataset^10^ is a dataset that focuses on the field of human activity recognition, employing the use of smartphones. The dataset comprises sensor data from accelerome-ters and gyroscopes of a Samsung Galaxy S3 smartphone. The activities recorded include walking, ascending stairs, descending stairs, sitting, standing, and lying down. The dataset is accessible via the UCI Machine Learning Repository. The UCI HAR Dataset is widely utilized due to its comprehensive sensor data and convenient accessibility.

The WISDM (Wireless Sensor Data Mining) Activity Prediction Dataset^48^ has been designed for the purpose of activity recognition using smartphone and smartwatch sensor data. The dataset includes accelerometer and gyroscope data for activities such as walking, jogging, sitting, standing, and climbing stairs. The dataset is accessible through the WISDM Project. The WISDM dataset is of benefit to research involving the use of mobile and wearable devices for monitoring human activity.

Another dataset of particular interest is BasicMotions,^47^ which encompasses a diverse range of movements, including walking, resting, running, and badminton. The dataset lacks specific metadata and is provided in Attribute-Relation File Format (ARFF). The dataset is accessible via Time Series Classification and comprises 40 datasets. The BasicMotions dataset is notable for its comprehensive coverage of various types of movements, rendering it an optimal choice for general studies on human movement recognition.

Two additional datasets warrant mention: Cricket^49^ and Racket Sports.^50^ These datasets include motion capture data, which can be used for specific applications, such as cricket, badminton, and squash. However, neither the exact number of test subjects nor records could be determined for either, nor any metadata was found for this study. Furthermore, RacketSports had no known publications in the context of these data.

In the study of IEMOCAP^22^, the emotional responses of the test subjects were recorded and subsequently analyzed, with fifty-three markers affixed to the subjects’ faces, and the minimal movement of various emotions was recorded.

The MotionSense dataset, introduced by Malekzadeh and colleagues,^11^ is designed for human activity and attribute recognition. It was collected using the accelerometer and gyroscope sensors of an iPhone 6s, which was placed in the front pocket of the participants during the experiments. The dataset includes time-series data recorded at a frequency of 50Hz and covers six different activities: walking downstairs, walking upstairs, walking, jogging, sitting, and standing. The MotionSense dataset has been used in various privacy and machine learning studies, particularly to explore how to protect sensitive data while maintaining the accuracy of activity recognition models. It’s freely available and has become a useful resource for HAR research.

The EmoPain dataset^12^ is a multimodal dataset designed to study pain-related behaviors, particularly in people with chronic lower back pain (CLBP). The dataset contains both healthy participants and individuals suffering from CLBP performing various physical tasks. It includes several types of data, such as body movement captured through motion capture (MoCap), surface electromyography (sEMG), and facial expression data collected during specific exercises.

The RealWorld HAR dataset,^13^ created by Sztyler and Stuckenschmidt, is designed for human activity recognition (HAR) in real-world environments. This dataset was collected using multiple wearable devices placed on different parts of the body, such as the head, chest, arms, wrists, and legs. It includes data from 15 participants performing 8 different activities: walking, running, standing, sitting, jumping, lying, and ascending or descending stairs. The RealWorld HAR dataset is widely used in studies related to mobile and wearable technology and is particularly valuable for tasks involving multi-modal sensor data and on-body localization of devices. The dataset is accessible and commonly used in machine learning benchmarks.

The HARTH (Human Activity Recognition Trondheim) dataset^14^ is designed for human activity recognition (HAR) in free-living environments. It was developed by researchers at the Norwegian University of Science and Technology (NTNU) and includes 22 participants who were recorded for 90 to 120 minutes during their regular working hours. The dataset aims to address limitations in previous HAR datasets by offering fixed sensor placements and high-quality annotations.

The HuGaDB (Human Gait Database)^27^ is a detailed dataset focused on human activity recognition and gait analysis, collected using a network of six wearable inertial sensors and electromyography (EMG) sensors. These sensors were placed on participants’ right and left thighs, shins, and feet, and two EMG sensors were placed on the quadriceps to measure muscle activity. The dataset is valuable for studying how different parts of the legs move during activities like walking, running, sitting, and taking stairs.

The HARSense dataset, developed by Nurul Amin Choudhury, Soumen Moulik, and Diptendu Sinha Roy,^15^ is designed for Human Activity Recognition (HAR) using smartphone sensor data. The dataset includes data from the accelerometers and gyroscopes of smartphones (such as the Poco X2 and Samsung Galaxy A32s), capturing six daily activities: walking, running, standing, sitting, going upstairs, and going downstairs. The smartphone sensors were mounted on the waist or placed in the front pockets of participants.

The ExtraSensory dataset, created by Yonatan Vaizman and colleagues,^16^ is a large-scale dataset designed for behavioral context recognition in real-world settings. The data was collected from 60 participants using smartphones and smartwatches over their regular daily activities. It includes data from multiple sensors such as accelerometers, gyroscopes, magnetometers, location, and audio. The dataset supports a wide range of human activity recognition tasks and offers a diverse set of labels related to daily and physical activities.

The KIT Whole-Body Human Motion Database, developed by Christian Mandery and colleagues,^17^ is a large-scale dataset designed for the analysis and synthesis of human motion. It captures detailed whole-body human motion using optical marker-based motion capture systems. The database is used for a variety of research applications, including human motion analysis, biomechanics, robotics, and rehabilitation. The database contains a wide range of motions, including daily activities, sports movements, and interactions with objects, all annotated for precise research. Researchers can also access metadata related to each motion, such as the subject’s physical attributes or the object involved in the motion.

The HDM05 dataset is a comprehensive motion capture (MoCap) dataset,^18^ designed to support research in areas like human motion analysis, biometrics, computer animation, and sports sciences. It contains more than three hours of systematically recorded motion capture data, amounting to 2,337 sequences spanning 130 motion classes, all performed by five actors. The data was captured using optical marker-based MoCap systems and is available in both C3D and ASF/AMC formats. The motions include a wide range of activities such as walking, running, jumping, dancing, sports movements, and object interactions.

The CMU Motion Capture Database^19^ is a widely used dataset in human motion analysis and computer animation. It contains thousands of motion sequences captured using an optical marker-based system, featuring a diverse range of activities such as walking, running, dancing, and sports movements. The data are freely available for research, but users are advised to handle noisy data in certain joints like the “toe” and “hand”. Each subject has a calibrated skeleton, and the dataset supports both academic and commercial research.

One of the most comprehensive datasets for motion capture is the Human3.6M, which was created by Catalin Ionescu and colleagues.^20^ The dataset comprises 3.6 million human poses captured using a high-speed motion capture system, with data recorded at 50 Hz. The dataset features 11 actors performing 17 different activities, including discussions, walking, running, and interacting with objects. It provides both 3D joint positions and high-resolution video data from multiple viewpoints, making it a key benchmark for 3D human pose estimation and action recognition models.

The FIT3D dataset, developed by Mihai Fieraru and colleagues,^21^ is a large-scale motion capture dataset designed for 3D human pose and motion analysis in fitness training. It contains over three million images and highly accurate 3D motion capture data, with corresponding human shape and skeleton configurations. The dataset focuses on 37 different exercises that cover all major muscle groups, performed by both fitness instructors and trainees. The motions were captured using multiple RGB cameras, creating a rich resource for analyzing exercise performance.

The 2D and 3D Full-Body Gesture Database, created by Bon-Woo Hwang, Sungmin Kim, and Seong-Whan Lee,^23^ is designed to capture and analyze daily human gestures. It includes both 2D video and 3D motion data of 14 representative daily gestures, performed by 20 subjects. These gestures were recorded using a combination of 12 sets of 3D motion cameras and 3 stereo cameras positioned at different angles to capture a complete view of each movement.

The HMDB (Human Motion Database),^24^ also known as HMDB51, is a large-scale video dataset designed for human action recognition. It consists of 6,766 video clips sourced from various places, including movies, public archives, and YouTube. The dataset is organized into 51 action categories, with each category containing at least 101 clips. These actions range from basic movements like walking, running, and jumping, to more complex activities involving object interactions, such as brushing hair, fencing, and dribbling.

The w-HAR dataset, created by Ganapati Bhat, Nicholas Tran, Holly Shill, and Umit Y. Ogras,^25^ is designed for human activity recognition (HAR) using low-power wearable devices. This dataset contains labeled data for seven activities (e.g., walking, jumping, sitting, standing) collected from 22 users. The data are gathered from two types of sensors: inertial measurement units (IMUs) and stretch sensors, providing both motion and stretch data for enhanced activity recognition.

The Smart-Insole Dataset^26^ is designed for gait analysis using pressure sensor insoles, with a particular focus on elderly individuals and Parkinson’s disease patients. The dataset includes data from 29 participants divided into three groups: healthy adults, elderly individuals, and Parkinson’s disease patients. The participants performed two key tests: the Walk Straight and Turn test and a modified version of the Timed Up and Go test.

The YARETA dataset^28^ focuses on asymptomatic human gait and general movements, including data from markers, inertial sensors, pressure insoles, and force plates. It was developed by Gautier Grouvel and his team at the University of Geneva. The dataset includes recordings from 10 participants who performed walking tasks and other movements such as squats and knee flexion/extension on a 10-meter walkway in a laboratory setting. The data provides 3D trajectories of 69 reflective markers, alongside acceleration and angular velocity data from inertial sensors, and pressure signals from insoles. The dataset is valuable for gait analysis and comparative studies between various motion capture systems.

The MAREA dataset (Movement Analysis in Real-world Environments using Accelerometers), developed by Siddhartha Khandelwal and Niclas Wickström,^29^ contains gait data from 20 healthy adults. The dataset captures walking and running activities in both indoor and outdoor environments using accelerometers placed on the waist, wrists, and ankles. It aims to evaluate the performance of gait event detection (GED) algorithms in real-world conditions, making it useful for analyzing movement variability across different terrains and speeds.

The NEWBEE dataset^30^ is a multi-modal gait database designed to capture natural everyday walking patterns in urban environments. It features recordings from 20 participants (5 females and 15 males, aged 18 to 69), using a full-body Lycra suit equipped with 17 IMU sensors, insole pressure sensors, and eye-tracking data. This rich dataset aims to facilitate research on natural gait patterns by providing data from a wide range of walking scenarios, including uneven terrains and varying speeds.

The MyPredict dataset, developed by Roessingh Research & Development,^31^ focuses on lower limb kinematics and electromyography (EMG) during gait-related activities. It was created to support activity prediction and human movement analysis, particularly for medical research and rehabilitation.

The Non-Laboratory Gait Dataset^32^ developed by Abhishek Sharma and colleagues focuses on capturing full-body kinematics and egocentric vision during human locomotion outside of controlled environments. This dataset was collected using inertial measurement units (IMUs) and includes activities such as walking, stair climbing, and obstacle course navigation.

The DU-MD (University of Dhaka Mobility Dataset)^33^ is a comprehensive sensor-based human action recognition (HAR) dataset developed for ubiquitous wearable devices. It con-tains data from 50 subjects who performed 10 different activities, including walking, sitting, standing, and falls. The data are collected using wrist-mounted sensors, primarily focusing on accelerometer data, making it ideal for studies related to healthcare monitoring, elderly surveillance, and fall detection.

The UniMiB SHAR dataset, developed by Daniela Micucci, Marco Mobilio, and Paolo Napoletano,^36^ is designed for human activity recognition (HAR) using data collected from smartphone accelerometers. The dataset consists of samples that cover a wide range of activities of daily living (ADL) and falls. Specifically, it includes nine types of ADLs (e.g., walking, sitting, running) and eight different types of falls (e.g., forward falls, backward falls, syncope). Data were collected from 30 subjects, mostly females (24 out of 30), aged between 18 and 60.

Overall, the systematic literature review demonstrates that the identified datasets cover a wide range of applications and sensor types. Most datasets offer detailed metadata and various data formats, facilitating their use for different research requirements. Platforms such as Figshare, Zenodo, and OSF play a crucial role in providing and facilitating access to these data. Some datasets, such as GaitRec and Hang-Time HAR, offer detailed metadata and specialized information, making them highly useful for specific studies. Others, such as the dataset by Senden and his colleagues as well as BasicMotions, have limitations in terms of metadata availability or specialization in certain movement types, which may restrict their applicability in certain research contexts. In conclusion, this systematic literature review provides a valuable resource for selecting appropriate datasets and facilitates building research in the field of human activity recognition on solid data foundations.

## 4 Discussion

In this section, we discuss the main findings from our systematic review in detail, emphasizing how challenges in data availability, standardization, and metadata variability affect current and future research in human activity recognition.

In this category, there was a notable increase in the difficulty of **data availability**. The data were either not always freely available or difficult to obtain when it was. In certain instances, data were accessible for download, but required the creation of an account with the website in question. Obviously, this research could only include sets that had available data, including those with prior registration.

It can be posited that the providers had disparate views on the manner in which their data should be made available. GitHub or similar sources remained an advantage.^25^ In the majority of cases, a download page was utilized to facilitate the download of a compressed folder in a ZIP format.

Moreover, there were considerable discrepancies between the **various movements tasks** The objective was to identify databases that define fundamental motions and contain a variety of these. However, only a multitude of incongruent databases or repositories containing different motions could be identified. These were included in the analysis specifically when, for example, different gait variations were presented.^27^ Nevertheless, a number of repositories containing different movement types were found to be convincing. However, these were largely comprised of simple movements such as standing, sitting, or lying in a variety of positions, with minimal variation. Further variation would be of great importance for the training of machine learning algorithms and is not represented here. It can be suggested that there is a plethora of movement data, yet it is limited to a narrow range of movements or small groups thereof.

**The metadata** of the datasets were divided into distinct categories. These included items such as information on the test subjects, the measurement equipment, or the measurement protocol. With the exception of a few cases, the subjects were typically provided only with information on their age, height, weight, or some other anthropometric data. Medical history or similar was rarely considered. Similarly, occupational history or origin was rarely explained. This further illustrates the paucity of information recorded about the subjects. Moreover, minimal information was provided regarding the measurement equipment and the measurement setup. Although it was possible to ascertain which equipment was employed in all datasets, no further details such as the type designation, recording frequency, or the placement of the sensors on the test subject were recorded. The measurement setup or measurement protocol was only mentioned in a few datasets, which makes it challenging to reproduce or understand the measurement. Furthermore, the number of subjects or recordings could not be consistently differentiated, which is why they are only labeled as “Number of datasets” in the table.

As previously indicated in the metadata section, the data pertaining to the **sensors type** are highly scalable. Additionally, the instrumentation employed for measurement is highly variable. In the majority of cases, a smartphone was employed as an inertial measurement unit (IMU) to capture the movement. Moreover, the majority of studies employed a single or a pair of sensors to conduct the measurement. This implies that it is not possible, for example, to identify correlations in IMU and EMG data derived from disparate movements. However, one of the most significant challenges was the **Data Type** or file format. Firstly, no database of any kind could be identified. The majority of websites use a file storage download system to facilitate the retrieval of voluminous data. Only a few sources provide a stream of the data, which then runs via GitHub or Figshare.^30^ Moreover, it was only possible to ascertain the file format or the structure of the data after downloading. The format of the measurement results was not adequately announced. It was not specified whether the data were presented in tabular form, with each column separated by a comma or similarly. Also, the data formats ranged from CSV over C3D to ARFF and similar ones, which made standardized analysis extremely challenging.

This systematic literature review is subject to several **limitations**. The study was limited to specific databases, which may have resulted in the omission of valuable datasets in lesser-known repositories. The search terms and criteria may have excluded relevant studies due to variations in terminology. The quality assessment focused on metadata and accessibility, without evaluating the actual quality of the data. The findings are temporally limited due to the continuous release of new datasets. The heterogeneity of dataset formats and metadata complicated data synthesis, affecting the consistency of conclusions. Furthermore, the review focused on English-language publications, introducing a potential language bias. Additionally, the emphasis on publicly accessible datasets excluded high-quality private datasets, limiting the understanding of the full data landscape. Future research should address these limitations to improve the robustness of reviews in biomechanical data, supporting better machine learning models and clinical applications.

To enhance the utility of biomechanical datasets for machine learning and AI applications, several critical improvements are needed. First, the development and adoption of standardized data formats and metadata protocols are essential. This will enable seamless integration of multiple datasets into machine learning frameworks and facilitate more effective data preprocessing and analysis. In addition, expanding both the volume and diversity of datasets is crucial. Many existing biomechanical datasets are limited to specific types of motion and lack a representation of varied demographics, which limits the generalizability of AI models. Increasing the scope of the dataset will improve the robustness of models in different populations and environments (Laboratory or Outside).

Incorporating multimodal data is a key step in advancing biomechanical databases. Instead of relying on a single source like MoCap or IMUs, combining data from MoCap, electromyography, wearable sensors, and force plates yields a more holistic view of human activity. MoCap excels in capturing kinematic details, while EMG reveals neuromuscular control, and wearables track daily-life movements. Used together, these data streams capture both the mechanical and physiological facets of movement.

From an SLR perspective, multimodal datasets expand the feature space for machine learning, enabling robust models that detect nuanced motor strategies or early signs of pathologies. However, these gains hinge on high-quality metadata detailing sensor configurations, sampling rates, participant demographics, and experimental conditions. Without comprehensive documentation, replicability suffers and the explanatory power of advanced analytics diminishes. Multimodal data enhances both model accuracy and interpretability. Sensor-fusion strategies can uncover subtle interdependencies among kinematics, muscle activation, and environmental factors. This leads to improved classification, earlier anomaly detection, and better trajectory forecastingcritical features for clinical applications, rehabilitation, and real-time monitoring.

Accessibility represents another crucial element in the promotion of extensive utilization and collaborative research endeavors. It is recommended that future datasets be made available through open-access platforms with centralized repositories. This would facilitate easy access for researchers and clinicians, enabling cross-study comparisons and accelerating advancements in the field. However, this must be balanced with stringent data protection regulations, such as the General Data Protection Regulation (GDPR), which impose strict guidelines on the collection, storage, and sharing of personal data.

To ensure compliance with the GDPR, datasets must be anonymized or de-identified wherever feasible, removing any personally identifiable information (PII) in order to safeguard the privacy of participants. Furthermore, it is essential to obtain explicit consent from participants, ensuring that they are fully aware of how their data will be utilized, shared, and stored. Furthermore, it is essential to implement comprehensive data governance frameworks that guarantee data security while providing the necessary transparency to participants. By establishing a balance between data accessibility and compliance with privacy regulations, future datasets can be made available in a manner that respects the rights of participants while simultaneously facilitating innovation and collaboration within the biomechanical research community.

Finally, there is a pressing need for the collection of real-world biomechanical data, as many existing datasets are derived from highly controlled environments. Capturing data in naturalistic settings such as during daily living activities will better reflect the complexities encountered in real-world applications, such as rehabilitation, sports performance monitoring, and fall detection. Such real-world datasets will enhance the ecological validity and applicability of machine learning models, allowing for more accurate predictions and improved outcomes in clinical and research contexts.

By systematically addressing these identified gaps, future biomechanical datasets can be optimized to support advanced artificial intelligence models more effectively. This enhancement will facilitate the development of improved methodologies for human movement analysis, rehabilitation strategies, and other applications within clinical settings.

## 5 Conclusion

This systematic literature review has revealed several critical challenges in leveraging opensource biomechanical datasets for AI in clinical biomechanics. Inconsistent metadata, nonstandardized data formats, and limited accessibility collectively undermine data reproducibility and hinder the performance of machine learning models. To overcome these challenges, it is imperative to establish comprehensive metadata standards that document acquisition protocols, sensor specifications, participant demographics, and experimental conditions. Such standards would ensure that datasets are fully interpretable and comparable across different studies. In addition, adopting uniform data formats and standardized nomenclature across the biomechanics community will facilitate the seamless integration of datasets, thereby enhancing data sharing and collaborative analysis. Furthermore, the promotion of centralized, open-access repositories with user-friendly access mechanisms and clear licensing agreements is essential to improve data availability and foster collaboration among researchers, clinicians, and industry stakeholders. Finally, coordinated multi-center trials that bring together expertise from academia, industry, and healthcare institutions are necessary to generate high-quality, interoperable datasets that can be reliably utilized for AI-driven clinical decision making. These integrated measures will not only enhance the quality and utility of biomechanical datasets but may also accelerate advancements in human movement analysis, injury prevention, and rehabilitation outcomes.

## Data Availability

All data produced in the present work are contained in the manuscript

**Daniel Friemert** is an academic at University of Applied Science in Remagen, specializing in data science, programming, mathematical methods, and motion analysis. He heads the Laboratory for Biomechanics, Ergonomics, and Virtual Reality, and directs the Institute for Sports Medical Technology and Sports Management. His research interests include computational biomechanics, dynamical systems, machine learning, and big data analytics in sports. Dr. Friemert holds a B.Sc. in Measurement and Sensor Technology and an M.Sc. in Applied Physics.

## Notes

### Competing Interest Statement

The authors have declared no competing interest.

### Funding Statement

This study did not receive any funding

## References

1 C. Z.-H. Ma, Z. Li, and C. He, “Advances in biomechanics-based motion analysis,” Bioengineering, vol. 10, no. 6, 2023. [Online]. Available: https://www.mdpi.com/2306-5354/10/6/677

2. “Exercise biomechanics for health: Evaluating lifelong activities for well-being,” https://www.ncbi.nlm.nih.gov/pmc/articles/PMC10048551/, (retrieved on: 26.06.2024).

3. “Impact of dataset size on deep learning model skill and performance estimates,” https://tinyurl.com/ybz2ylvd, (retrieved on: 26.06.2024).

4 A. P. Siddaway, A. M. Wood, and L. V. Hedges, “How to do a systematic review: A best practice guide for conducting and reporting narrative reviews, meta-analyses, and meta-syntheses,” Annual Review of Psychology, vol. 70, no. Volume 70, 2019, pp. 747–770, 2019. [Online]. Available: https://www.annualreviews.org/content/journals/ 10.1146/annurev-psych-010418-102803

5. C. Wohlin, “Guidelines for snowballing in systematic literature studies and a replication in software engineering,” in Proceedings of the 18th International Conference on Evaluation and Assessment in Software Engineering, ser. EASE ’14. New York, NY, USA: Association for Computing Machinery, 2014. [Online]. Available: 10.1145/2601248.2601268

6 B. Horsak, D. Slijepcevic, A. M. Raberger, C. Schwab, M. Worisch, and M. Zeppelzauer, “GaiTRec, a large-scale ground reaction force dataset of healthy and impaired gait,” Scientific Data, vol. 7, no. 1, p. 143, 2020.

7 A. Hoelzemann, J. L. Romero, M. Bock, K. V. Laerhoven, and Q. Lv, “Hang-time har: A benchmark dataset for basketball activity recognition using wrist-worn inertial sensors,” Sensors, vol. 23, no. 13, 2023. [Online]. Available: https://www.mdpi.com/1424-8220/23/13/5879

8 R. Senden, R. Marcellis, K. Meijer, P. Willems, T. Lenssen, H. Staal, Y. Janssen, V. Groen, R. J. Vermeulen, and M. Witlox, “Dataset of 3d gait analysis in typically developing children walking at three different speeds on an instrumented treadmill in virtual reality,” Data in Brief, vol. 48, p. 109142, 2023. [Online]. Available: https://www.sciencedirect.com/science/article/pii/S2352340923002615

9. T. Le Nguyen, S. Gsponer, I. Ilie, M. O’Reilly, and G. Ifrim, “Interpretable time series classification using linear models and multi-resolution multi-domain symbolic representations,” 05 2020.

10 J. e. a. Reyes-Ortiz, “Human Activity Recognition Using Smartphones,” UCI Machine Learning Repository, 2012, DOI: 10.24432/C54S4K.

11. M. Malekzadeh, R. G. Clegg, A. Cavallaro, and H. Haddadi, “Mobile sensor data anonymization,” in Proceedings of the International Conference on Internet of Things Design and Implementation, ser. IoTDI ’19. New York, NY, USA: Association for Computing Machinery, 2019, p. 4958. [Online]. Available: 10.1145/3302505.3310068

12 P. Lucey, J. F. Cohn, K. M. Prkachin, P. E. Solomon, and I. Matthews, “Painful data: The unbc-mcmaster shoulder pain expression archive database,” in 2011 IEEE International Conference on Automatic Face & Gesture Recognition (FG), 2011, pp. 57–64.

13 T. Sztyler and H. Stuckenschmidt, “On-body localization of wearable devices: An investigation of position-aware activity recognition,” in 2016 IEEE International Conference on Pervasive Computing and Communications (PerCom), 2016, pp. 1–9.

14 A. Logacjov, K. Bach, A. Kongsvold, H. B. Bårdstu, and P. J. Mork, “Harth: A human activity recognition dataset for machine learning,” Sensors, vol. 21, no. 23, 2021. [Online]. Available: https://www.mdpi.com/1424-8220/21/23/7853

15 N. A. Choudhury, S. Moulik, and D. S. Roy, “Harsense: Statistical human activity recognition dataset,” 2021. [Online]. Available: 10.21227/9pt3-2m34

16 Y. Vaizman, K. Ellis, and G. Lanckriet, “Recognizing detailed human context in the wild from smartphones and smartwatches,” IEEE Pervasive Computing, vol. 16, no. 4, pp. 62–74, 2017.

17 C. Mandery, Terlemez, M. Do, N. Vahrenkamp, and T. Asfour, “Unifying representations and large-scale whole-body motion databases for studying human motion,” IEEE Transactions on Robotics, vol. 32, no. 4, pp. 796–809, 2016.

18. M. Müller, T. Röder, M. Clausen, B. Eberhardt, B. Krüger, and A. Weber, “Documentation mocap database hdm05,” Universität Bonn, Tech. Rep. CG-2007-2, June 2007.

19. M. Balazia and P. Sojka, An Evaluation Framework and Database for MoCap-Based Gait Recognition Methods. Springer International Publishing, 2017, p. 3347. [Online]. Available: 10.1007/978-3-319-56414-2_3

20 C. Ionescu, D. Papava, V. Olaru, and C. Sminchisescu, “Human3.6m: Large scale datasets and predictive methods for 3d human sensing in natural environments,” IEEE Transactions on Pattern Analysis and Machine Intelligence, vol. 36, no. 7, pp. 1325–1339, 2014.

21 M. Fieraru, M. Zanfir, S. C. Pirlea, V. Olaru, and C. Sminchisescu, “Aifit: Automatic 3d human-interpretable feedback models for fitness training,” in 2021 IEEE/CVF Conference on Computer Vision and Pattern Recognition (CVPR), 2021, pp. 9914–9923.

22 C. Busso, M. Bulut, C.-C. Lee, A. Kazemzadeh, E. Mower Provost, S. Kim, J. Chang, S. Lee, and S. Narayanan, “Iemocap: Interactive emotional dyadic motion capture database,” Language Resources and Evaluation, vol. 42, pp. 335–359, 12 2008.

23 B.-W. Hwang, S. Kim, and S.-W. Lee, “A full-body gesture database for human gesture analysis,” International Journal of Pattern Recognition and Artificial Intelligence, vol. 21, no. 06, pp. 1069–1084, 2007. [Online]. Available: 10.1142/S0218001407005806

24 H. Kuehne, H. Jhuang, E. Garrote, T. Poggio, and T. Serre, “Hmdb: A large video database for human motion recognition,” in 2011 International Conference on Computer Vision, 2011, pp. 2556–2563.

25 G. Bhat, N. Tran, H. Shill, and U. Y. Ogras, “w-har: An activity recognition dataset and framework using low-power wearable devices,” Sensors, vol. 20, no. 18, 2020. [Online]. Available: https://www.mdpi.com/1424-8220/20/18/5356

26 C. Chatzaki, V. Skaramagkas, N. Tachos, G. Christodoulakis, E. Maniadi, Z. Kefalopoulou, D. I. Fotiadis, and M. Tsiknakis, “The smart-insole dataset: Gait analysis using wearable sensors with a focus on elderly and parkinsons patients,” Sensors, vol. 21, no. 8, 2021. [Online]. Available: https://www.mdpi.com/1424-8220/21/8/2821

27. R. Chereshnev and A. Kertész-Farkas, “Hugadb: Human gait database for activity recognition from wearable inertial sensor networks,” in Analysis of Images, Social Networks and Texts, W. M. van der Aalst, D. I. Ignatov, M. Khachay, S. O. Kuznetsov, V. Lempitsky, I. A. Lomazova, N. Loukachevitch, A. Napoli, A. Panchenko, P. M. Pardalos, A. V. Savchenko, and S. Wasserman, Eds. Cham: Springer International Publishing, 2018, pp. 131–141.

28 G. Grouvel, L. Carcreff, F. Moissenet, and S. Armand, “A dataset of asymptomatic human gait and movements obtained from markers, imus, insoles and force plates,” Scientific Data, vol. 10, 03 2023.

29 S. Khandelwal and N. Wickström, “Evaluation of the performance of accelerometer-based gait event detection algorithms in different real-world scenarios using the marea gait database,” Gait & Posture, vol. 51, pp. 84–90, 2017. [Online]. Available: https://www.sciencedirect.com/science/article/pii/S0966636216305859

30 V. Losing and M. Hasenjäger, “A multi-modal gait database of natural everyday-walk in an urban environment,” Scientific Data, vol. 9, 08 2022.

31 R. Schulte, E. Prinsen, L. Schaake, R. Paassen, M. Zondag, E. Staveren, M. Poel, and J. Buurke, “Database of lower limb kinematics and electromyography during gait-related activities in able-bodied subjects,” Scientific Data, vol. 10, 07 2023.

32 A. Sharma, V. Rai, M. Calvert, Z. Dai, Z. Guo, D. Boe, and E. Rombokas, “A non-laboratory gait dataset of full body kinematics and egocentric vision,” Scientific Data, vol. 10, 01 2023.

33 S. S. Saha, S. Rahman, M. J. Rasna, A. Mahfuzul Islam, and M. A. Rahman Ahad, “Dumd: An open-source human action dataset for ubiquitous wearable sensors,” in 2018 Joint 7th International Conference on Informatics, Electronics & Vision (ICIEV) and 2018 2nd International Conference on Imaging, Vision & Pattern Recognition (icIVPR), 2018, pp. 567–572.

34 M. Shoaib, S. Bosch, O. D. Incel, H. Scholten, and P. J. M. Havinga, “Fusion of smartphone motion sensors for physical activity recognition,” Sensors, vol. 14, no. 6, pp. 10 146–10 176, 2014. [Online]. Available: https://www.mdpi.com/1424-8220/14/6/10146

35 W. Ugulino, D. Cardador, K. Vega, E. Velloso, R. Milidiú, and H. Fuks, “Wearable computing: Accelerometers’ data classification of body postures and movements,” vol. 7589, 10 2012.

36 D. Micucci, M. Mobilio, and P. Napoletano, “Unimib shar: A dataset for human activity recognition using acceleration data from smartphones,” Applied Sciences, vol. 7, no. 10, 2017. [Online]. Available: https://www.mdpi.com/2076-3417/7/10/1101

37. M. Zhang and A. A. Sawchuk, “Usc-had: a daily activity dataset for ubiquitous activity recognition using wearable sensors,” in Proceedings of the 2012 ACM Conference on Ubiquitous Computing, ser. UbiComp ’12. New York, NY, USA: Association for Computing Machinery, 2012, p. 10361043. [Online]. Available: 10.1145/2370216.2370438

38. G. Weiss, “WISDM Smartphone and Smartwatch Activity and Biometrics Dataset,” UCI Machine Learning Repository, 2019, DOI: 10.24432/C5HK59.

39. T. Huynh, M. Fritz, and B. Schiele, “Discovery of activity patterns using topic models,” in Proceedings of the 10th International Conference on Ubiquitous Computing, ser. UbiComp ’08. New York, NY, USA: Association for Computing Machinery, 2008, p. 1019. [Online]. Available: 10.1145/1409635.1409638

40 J. R. Kwapisz, G. M. Weiss, and S. A. Moore, “Activity recognition using cell phone accelerometers,” SIGKDD Explor. Newsl., vol. 12, no. 2, p. 7482, mar 2011. [Online]. Available: 10.1145/1964897.1964918

41 A. Yang, R. Jafari, S. Sastry, and R. Bajcsy, “Distributed recognition of human actions using wearable motion sensor networks,” JAISE, vol. 1, pp. 103–115, 01 2009.

42 A. Reiss and D. Stricker, “Introducing a new benchmarked dataset for activity monitoring,” in 2012 16th International Symposium on Wearable Computers, 2012, pp. 108–109.

43. N. Kawaguchi, N. Ogawa, Y. Iwasaki, K. Kaji, T. Terada, K. Murao, S. Inoue, Y. Kawahara, Y. Sumi, and N. Nishio, “Hasc challenge: gathering large scale human activity corpus for the real-world activity understandings,” in Proceedings of the 2nd Augmented Human International Conference, ser. AH ’11. New York, NY, USA: Association for Computing Machinery, 2011. [Online]. Available: 10.1145/1959826.1959853

44. S. S. Intille, K. Larson, J. S. Beaudin, J. Nawyn, E. M. Tapia, and P. Kaushik, “A living laboratory for the design and evaluation of ubiquitous computing technologies,” in CHI ’05 Extended Abstracts on Human Factors in Computing Systems, ser. CHI EA ’05. New York, NY, USA: Association for Computing Machinery, 2005, p. 19411944. [Online]. Available: 10.1145/1056808.1057062

45 D. Roggen, A. Calatroni, M. Rossi, T. Holleczek, K. Förster, G. Tröster, P. Lukowicz, D. Bannach, G. Pirkl, A. Ferscha, J. Doppler, C. Holzmann, M. Kurz, G. Holl, R. Chavarriaga, H. Sagha, H. Bayati, M. Creatura, and J. d. R. Millàn, “Collecting complex activity datasets in highly rich networked sensor environments,” in 2010 Seventh International Conference on Networked Sensing Systems (INSS), 2010, pp. 233–240.

46. O. Baños, M. Damas, H. Pomares, I. Rojas, M. A. Tóth, and O. Amft, “A benchmark dataset to evaluate sensor displacement in activity recognition,” in Proceedings of the 2012 ACM Conference on Ubiquitous Computing, ser. UbiComp ’12. New York, NY, USA: Association for Computing Machinery, 2012, p. 10261035. [Online]. Available: 10.1145/2370216.2370437

47. “Dataset: Basicmotions time series classification,” https://timeseriesclassification.com/description.php?Dataset=BasicMotions, (retrieved on: 15.06.2024).

48. “Wisdm smartphone and smartwatch activity and biometrics dataset,” https://www.cis.fordham.edu/wisdm/dataset.php, (retrieved on: 19.06.2024).

49. “Dataset: Cricket time series classification,” https://www.timeseriesclassification.com/description.php?Dataset=Cricket, (retrieved on: 20.06.2024).

50. “Dataset: Racketsports time series classification,” https://www.timeseriesclassification.com/description.php?Dataset=RacketSports, (retrieved on: 19.06.2024).

